# Evaluate epidemiologic burden, treatment patterns and treatment outcomes of advanced prostate cancer in Hong Kong using territory-wide electronic medical records

**DOI:** 10.64898/2026.10.12.26365031

**Authors:** Yuanshi Jiao, Jiaqi Wang, Xue Li

## Abstract

**Background:** Prostate cancer (PCa) is the most common urological malignancy globally and remains a major cause of cancer mortality among men. In Hong Kong, a comprehensive evaluation of disease burden, real-world treatment patterns, and outcomes across clinically relevant PCa subgroups is essential for identifying unmet needs and optimising care delivery.

**Objectives:** This study aims to characterise the burden of PCa and patterns of care in Hong Kong by (1) estimating annual patient load (2) describing treatment patterns, and (3) evaluating real-world outcomes among patients receiving systemic therapy (chemotherapy or novel hormonal agents [NHA]) and novel agents (immunotherapy, poly ADP-ribose polymerase [PARP] inhibitors, radiation therapy).

**Methods:** We conducted a population-based retrospective cohort study using electronic medical records from the Hong Kong Hospital Authority (HA). The cohort included patients diagnosed with PCa between January 1, 2012 and December 31, 2024. Subgroup analyses included patients with metastatic hormone-sensitive prostate cancer (mHSPC) and metastatic castration-resistant prostate cancer (mCRPC) and were further stratified by the treatment received. Descriptive analyses summarised annual patient load and treatment patterns over 2018-2022. We evaluated treatment outcomes from 2012 to 2024, including overall survival, progression-free survival, number of hospitalisations, and main adverse events.

**Results:** From 2018 to 2022, prevalence increased for prostate cancer (12,014 to 15,610), mHSPC (1,188 to 1,779), and mCRPC (167 to 290), while annual mCRPC incidence rose from 125 to 274. There was an increasing number of prevalent and new users of the investigated novel treatment, including first-/second-/third-line therapy for mCRPC. Compared with patients treated with NHAs, chemotherapy showed a numerically higher rate of hospitalisations, bone treatment initiation and higher hematologic toxicity.

**Conclusions:** This population-based retrospective study provides Hong Kong–specific evidence on PCa burden, real-world management, and outcomes across key subgroups, thereby supporting policy planning, formulary evaluation, and targeted strategies to address unmet clinical needs.

**Funding:** This work was supported by a grant from Novartis.

## INTRODUCTION

Prostate cancer is among the most common malignancies in men worldwide and is an increasingly important public health concern in the Asia–Pacific region. Historically, prostate cancer incidence in Asian populations has been substantially lower than in Western countries; however, this gap has progressively narrowed^1^. With population ageing and the wider uptake of prostate-specific antigen (PSA) testing, incidence has risen over time in settings such as Hong Kong.^2^

In parallel, the systemic treatment of advanced prostate cancer has changed markedly over the past decade. Androgen deprivation therapy (ADT) monotherapy was long the foundation of care, but multiple landmark trials have established treatment intensification as the contemporary standard. Adding novel hormonal agents (NHAs)^3–5^ and/or taxane-based chemotherapy^6^ to continuous ADT improves overall survival and is now recommended in international guidelines. Therapeutic options have also expanded in metastatic castration-resistant prostate cancer (mCRPC) to include targeted agents, such as radium-223 for patients with bone-predominant metastases^7^, and poly(ADP-ribose) polymerase (PARP) inhibitors (e.g., olaparib) for those with homologous recombination repair (HRR) gene alterations^8^.

As the therapeutic armamentarium expands, a contemporary assessment of epidemiologic burden, treatment burden, and actual care pathways is needed to quantify how different disease stages contribute to population-level burden over time, anticipate service and resource needs, evaluate equity and access, and support local guideline implementation. This is particularly relevant in Hong Kong, where prostate cancer is now the third most common cancer in men.^9^ However, evidence describing contemporary treatment pathways and real-world uptake of these therapies remains limited.

To address this gap, we used a comprehensive, territory-wide electronic medical record database to quantify the epidemiologic burden of prostate cancer, characterise evolving treatment patterns, and evaluate real-world outcomes for men with advanced disease in Hong Kong.

## METHOD

### Study Design and Data Source

This study employed a population-based retrospective cohort design utilising electronic medical records (EMRs) from the Hong Kong Hospital Authority (HA). The HA is the primary provider of publicly funded healthcare in Hong Kong, serving over 7.3 million residents—predominantly of Asian descent—and accounting for approximately 80% of all hospital admissions in the territory^10^. De-identified, unique patient reference numbers were utilised to extract comprehensive longitudinal data, including diagnostic records, prescription histories, healthcare utilisation, and mortality outcomes.

### Study Population and Subgroup Definitions

The target population comprised patients diagnosed with prostate cancer, identified via the International Classification of Diseases, Ninth Revision, Clinical Modification (ICD-9-CM) code 185, between January 1, 2012, and December 31, 2024. Patients were further stratified into clinical subgroups based on disease status. Metastatic disease was defined by the presence of subsequent malignant neoplasms using ICD-9-CM codes 196–199. Castration-resistant prostate cancer (CRPC) was identified using literature-reported criteria, characterised by castrate serum testosterone levels (< 50 ng/dL or 1.7 nmol/L) concurrent with biochemical progression (defined as a PSA increase of > 2 ng/mL and a 50% increase over the nadir)^11^. Patients presenting with a metastatic diagnosis without a prior CRPC identification were classified as having metastatic hormone-sensitive prostate cancer (mHSPC).

### Treatment Exposures and Sequencing

To evaluate evolving treatment patterns, systemic therapies were categorised into distinct clusters: chemotherapy (docetaxel, cabazitaxel), NHAs / androgen receptor pathway inhibitors (abiraterone, enzalutamide, apalutamide, darolutamide), radiopharmaceuticals (radium-223), PARP inhibitors (olaparib), and immunotherapy (pembrolizumab). The first therapeutic dispensing record following an mHSPC or mCRPC diagnosis denoted the initiation of first-line therapy. The commencement of a drug from a different treatment cluster marked the transitionv to a subsequent line of therapy. Patients were grouped annually by dispensing dates to quantify both prevalent (continuing) and incident (newly initiated) users.

### Outcome Measures

The study evaluated outcomes across three primary domains:Epidemiological Burden: The annual incidence and prevalence of overall prostate cancer, mHSPC, and mCRPC were calculated from 2018 to 2022. Prevalence was measured at the 1^st^ July of each calendar year, while incidence captured new cases within that year. 2) Clinical Efficacy: Real-world treatment outcomes were assessed by measuring median overall survival (OS), 2-year OS rates, median PSA-progression-free survival (PFS), and 2-year PSA-PFS rates. Patient follow-up started from the initiation of treatment to the earliest date of death or administrative censoring (Dec 31, 2024). Patients were grouped by the first exposure after the corresponding disease was diagnosed. Treatment switches were not considered during follow-up. 3) Treatment patterns: the first date of drug dispensing for certain predefined categories was the start of a certain line of treatment, and subsequent records of treatment in the same categories were dismissed (no in-category switch back was considered) and retained in separate Sankey diagrams. Patients without any investigated events, including death and initiation of any line of treatment, were excluded from the Sankey diagram. 4) Treatment Burden and Safety: Healthcare resource utilisation was quantified using median annualised inpatient admissions and lengths of stay (LOS). Safety and tolerability were assessed by tracking the incidence of Common Terminology Criteria for Adverse Events (CTCAE) Grade 2 and 3 bone marrow-related toxicities (anaemia, neutropenia, and thrombocytopenia) and the proportion of patients requiring bone marrow supportive treatments (filgrastim, pegfilgrastim, or blood transfusions).

### Statistical Analysis

Descriptive analyses were conducted to summarise all epidemiological treatment, and clinical outcomes. Subgroup analyses were explored based on specific treatment categories and lines of therapy. All statistical procedures were performed using R software, version 4.4.2 (R Foundation for Statistical Computing, Vienna, Austria), and the results were cross-checked by two independent investigators (YJ and JW) to ensure accuracy.

## RESULTS

### Epidemiologic Burden

Between 2018 and 2022, a progressive increase in the territory-wide epidemiologic burden of prostate cancer was observed in Hong Kong. The annual prevalence of overall prostate cancer rose steadily from 12,403 cases in 2018 to 15,925 cases in 2022. Parallel upward trends were noted in advanced stages; the prevalence of metastatic prostate cancer increased from 1,389 to 2,136 cases, while mHSPC prevalence grew from 1,215 to 1,811 cases over the same period. Furthermore, the annual incidence of mCRPC more than doubled, escalating from 125 new cases in 2018 to 274 new cases in 2022. (**Table 1**).

**Table 1.**
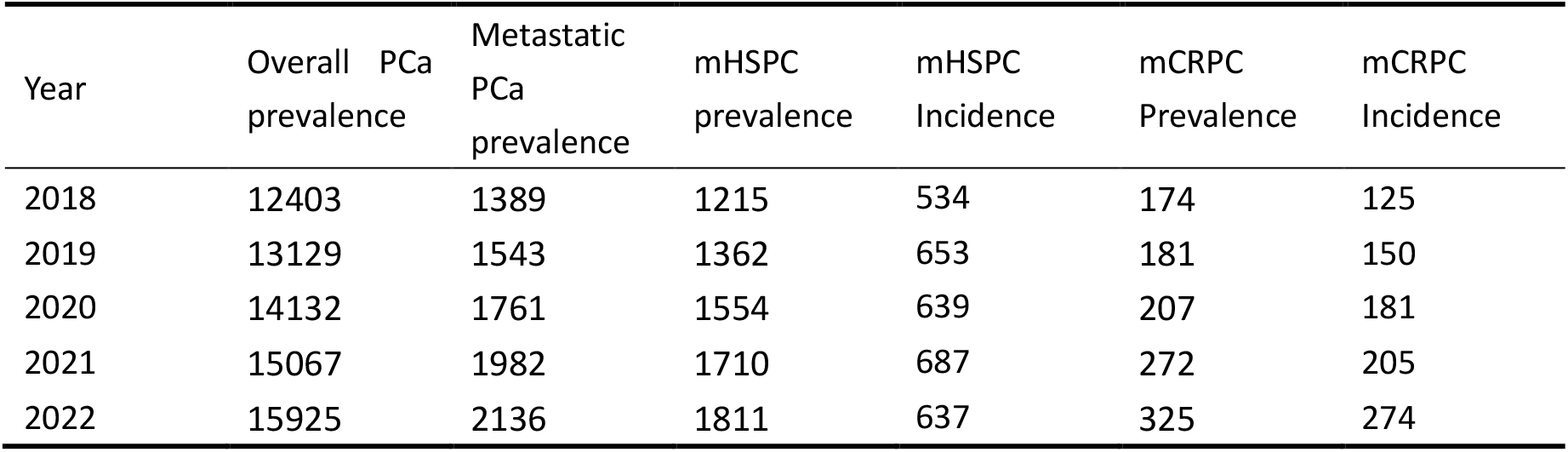
Epidemiological Trends of Prostate Cancer in Hong Kong (2018–2022) Prevalence was measured by the mid-year cohort. Abbreviation: PCa-Prostate cancer; mHSPC-metastatic hormone-sensitive prostate cancer; mCRPC-metastatic castration-resistant prostate cancer

### Real-World Treatment Patterns and Sequencing

Analysis of systemic therapeutic pathways revealed a substantial expansion in the active volume of both incident and prevalent treatment users. In the mCRPC setting, new users initiating first-line systemic therapy grew from 36 in 2018 to 98 in 2022. Sequential NHA use was also prevalent, with more patients switching from enzalutamide to abiraterone over time (**Table 2**). Additionally, the median overall duration of therapy prolonged significantly from 17.84 months in 2018 to 26.86 months in 2022. **Figure 1** presents the treatment trajectory after mHSPC diagnosis. Abiraterone, enzalutamide, and docetaxel consisted of the three dominant treatment options after mHSPC diagnosis, and relatively few patients have access to immunotherapy, PARP inhibitors throughout their treatment journey, or radiopharmaceuticals in mCRPC setting. There were 5876 patients (68%) who did not receive any investigated therapies after mHSPC diagnosis in our analysis.

**Table 2.** Key Treatment Inception, Sequencing Patterns, and Line Progression (2018–2022) Abbreviation: mHSPC-metastatic hormone-sensitive prostate cancer; mCRPC-metastatic castration-resistant prostate cancer; NHA-novel hormonal agent; IQR-interquartile range; PCa-prostate cancer *Line of investigated treatment starting from mHSPC including chemotherapy, NHA, Radium-223, PARP, Immunotherapy, not including class-switch back

| Treatment Subgroup & Patient Status | 2018 | 2019 | 2020 | 2021 | 2022 |
| --- | --- | --- | --- | --- | --- |
| Number of mCRPC patients stay with first-line treatment* | 93 | 123 | 157 | 211 | 271 |
| Number of mCRPC patients initiated first-line treatment* | 36 | 57 | 63 | 74 | 98 |
| Number of mCRPC patients stay with second-line treatment* | 65 | 92 | 98 | 128 | 168 |
| Number of mCRPC patients initiated second-line treatment* | 36 | 46 | 44 | 62 | 72 |
| Number of mCRPC patients stay with third-line treatment* | 16 | 23 | 30 | 36 | 37 |
| Number of mCRPC patients initiated third-line treatment* | 12 | 16 | 20 | 17 | 20 |
| Treatment duration (days) of NHA (Median [IQR]) | 154 (240) | 159 (222) | 168 (197) | 200 (299) | 229 (311) |
| Treatment duration (days) of chemotherapy (Median [IQR]) | 112 (74) | 114 (123) | 127 (116) | 133 (96) | 137 (103) |
| mHSPC NHA: Incident Users | 79 | 100 | 136 | 154 | 150 |
| mHSPC Chemotherapy: Incident Users | 26 | 54 | 53 | 55 | 45 |
| Sequential NHA after PCa (Abiraterone → Enzalutamide): Incident Users | 34 | 28 | 26 | 24 | 26 |
| Sequential NHA after PCa (Enzalutamide → Abiraterone): Incident Users | 10 | 20 | 20 | 25 | 25 |
| Sequential treatment after metastasis (Chemo → NHA): Incident Users | 51 | 55 | 64 | 89 | 101 |
| Sequential treatment after metastasis (NHA → Chemo): Incident Users | 11 | 30 | 29 | 49 | 50 |
| Both NHA and Chemotherapy after metastasis: Prevalence Users | 125 | 180 | 224 | 322 | 372 |

**Figure 1.**
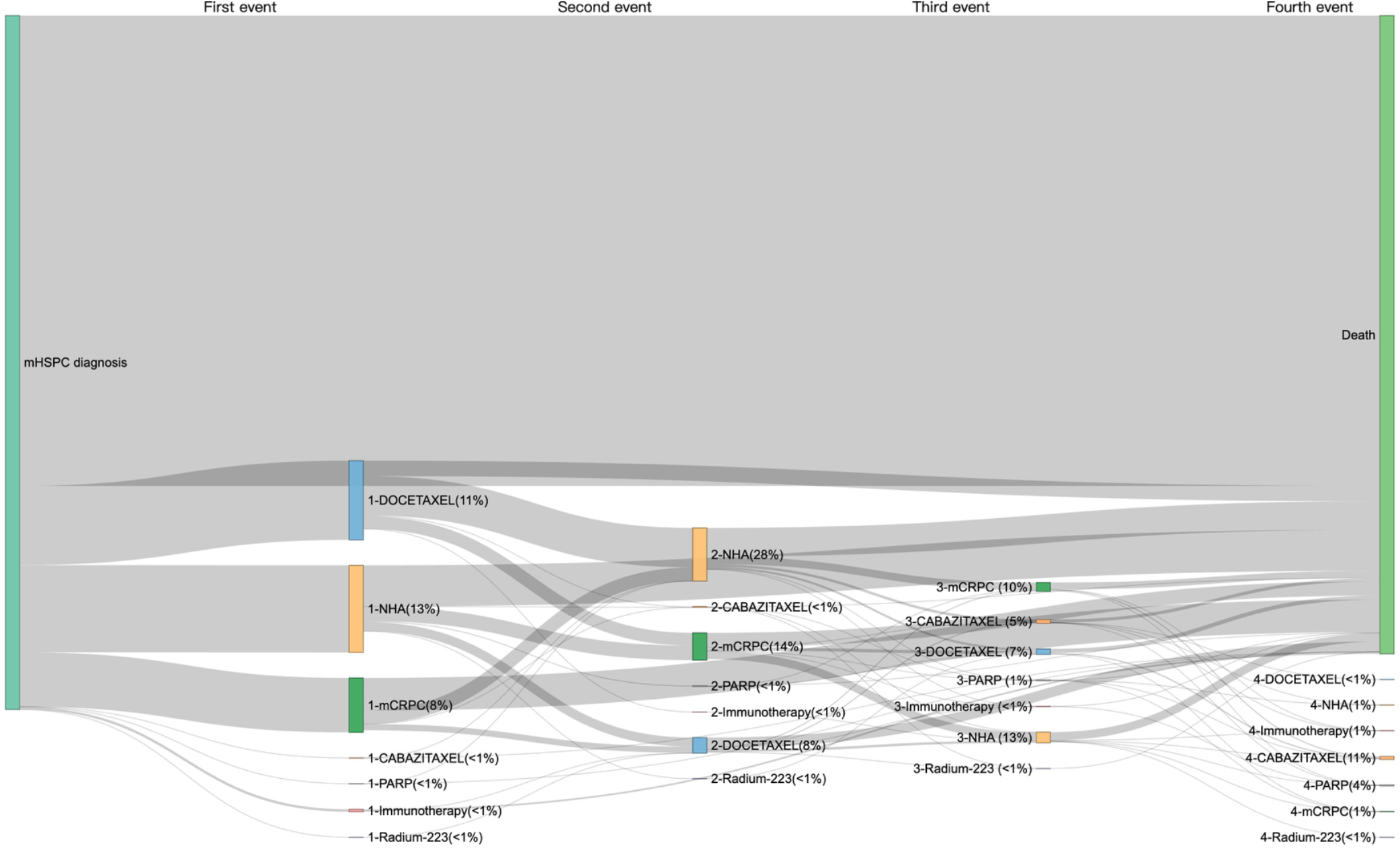
Treatment trajectory and disease status of mHSPC patients (2018-2022) Abbreviation: mHSPC-metastatic hormone-sensitive prostate cancer; NHA-noval hormonal agent; mCRPC-metastatic castration-resistant prostate cancer; PARP-poly(ADP-ribose) polymerase

**Figure 2.**
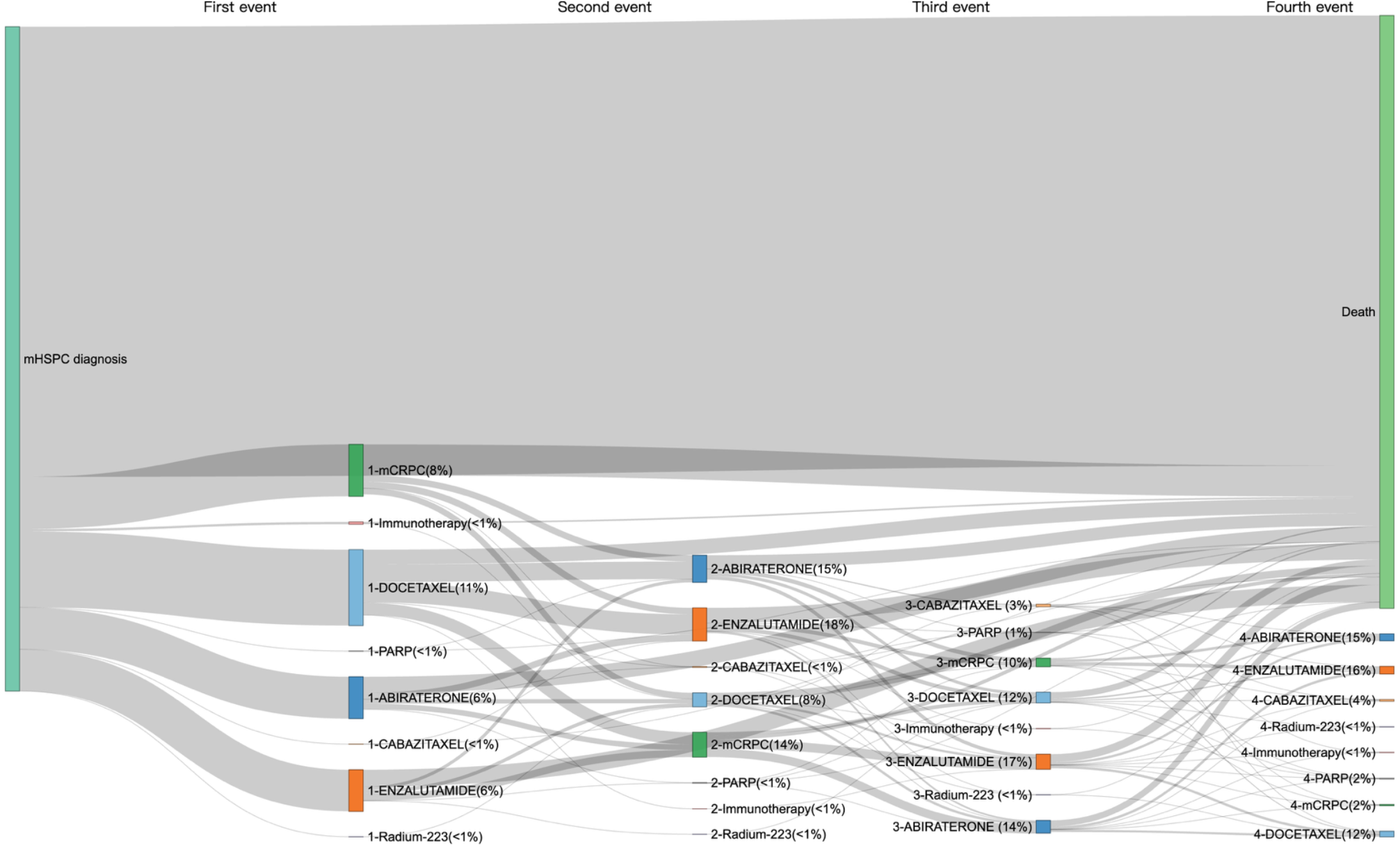
Treatment trajectory and disease status of mHSPC patients (2018-2022) with in-category switch-back Abbreviation: mHSPC-metastatic hormone-sensitive prostate cancer; mCRPC-metastatic castration-resistant prostate cancer; PARP-poly(ADP-ribose) polymerase

### Clinical Outcomes and Healthcare Resource Utilisation

In the aggregated cohort (2012–2024), real-world treatment outcomes varied by clinical stage and drug class (**Table 3** and **Table 4**). Among the mHSPC cohort, patients treated with chemotherapy (N=1,137) experienced a numerically longer median overall survival (OS) of 29.2 months compared to 20.3 months for those receiving NHAs (N=1,402). Median PSA-PFS was 12.0 months in the chemotherapy arm versus 10.0 months in the NHA arm. However, chemotherapy was tied to higher healthcare utilisation, yielding a median annualised hospitalisation frequency of 7.12 admissions compared to 4.38 for NHAs. Among the mCRPC cohort, survival time was reduced across both cohorts in the advanced castration-resistant stage. The median OS was 14.4 months for chemotherapy (N=207) and 13.3 months for NHAs (N=630). The chemotherapy cohort showed a median PFS of 6.4 months compared to 4.6 months for NHAs. Annualised inpatient stays were extensive for both groups, reaching a median LOS of 35.4 days for NHAs and 37.36 days for chemotherapy.

**Table 3.** Real-World Clinical and Healthcare Outcomes in the metastatic hormone-sensitive prostate cancer cohort. *Treatment switch refer to patients switch to the counterparts (i.e. Novel hormonal agents or chemotherapy).

| Outcomes | Novel Hormonal Agents (N = 1,402) | Chemotherapy (N = 1,137) |
| --- | --- | --- |
| Treatment switch* during follow-up | 202 (14.4%) | 664 (58.4%) |
| Clinical Efficacy Outcomes |  |  |
| Median Overall Survival (Interquartile Range) | 20.3 (18.7-21.9) months | 29.2 (26.7-32.1) months |
| 2-Year Overall Survival Rate | 44.7% (42.1%-47.5%) | 57.1% (54.2%-60.1%) |
| Median Progression-Free Survival (Interquartile Range) | 10.0 (8.9-10.8) months | 12.0 (11.2-13.3) months |
| 2-Year Progression-Free Survival Rate | 30.3% (27.9%-33.0%) | 33.1% (30.4%-36.0%) |
| Healthcare Resource Utilization |  |  |
| Median Annualised Hospitalization Frequency (times) | 4.38 | 7.12 |
| Median Annualised Length of Stay (days) | 22.74 | 22.24 |
| Safety & Hematologic Adverse Events |  |  |
| Bone Marrow Supportive Treatment Required | 18.12% | 56.11% |
| Anemia Grade 2 | 68.19% | 72.56% |
| Anemia Grade 3 | 48.43% | 52.68% |
| Neutropenia Grade 2 | 8.92% | 25.33% |
| Neutropenia Grade 3 | 4.56% | 23.22% |
| Thrombocytopenia Grade 2 | 18.54% | 21.90% |
| Thrombocytopenia Grade 3 | 12.84% | 16.62% |

**Table 4.** Real-World Clinical and Healthcare Outcomes in the metastatic castration-resistance cancer cohort. *Treatment switch refer to patients switch to the counterparts (i.e. Novel hormonal agents or chemotherapy).

| Outcomes | Novel Hormonal Agents (N = 630) | Chemotherapy (N = 207) |
| --- | --- | --- |
| Treatment switch* during follow-up | 127 (20.2%) | 91 (44%) |
| Clinical Efficacy Outcomes |  |  |
| Median Overall Survival (Interquartile Range) | 13.3 (12.0-14.9) months | 14.4 (12.3-17.3) months |
| 2-Year Overall Survival Rate | 28.0% (24.6%-31.9%) | 26.5% (21.0%-33.5%) |
| Median Progression-Free Survival (Interquartile Range) | 4.6 (4.3-5.1) months | 6.4 (5.7-7.2) months |
| 2-Year Progression-Free Survival Rate | 12.5% (10.0%-15.5%) | 9.0% (5.7%-14.2%) |
| Healthcare Resource Utilization |  |  |
| Median Annualised Hospitalization Frequency (times) | 6.24 | 9.17 |
| Median Annualised Length of Stay (days) | 35.40 | 37.36 |
| Safety & Hematologic Adverse Events |  |  |
| Bone Marrow Supportive Treatment Required | 27.46% | 53.14% |
| Anemia Grade 2 | 80.32% | 82.13% |
| Anemia Grade 3 | 61.11% | 60.39% |
| Neutropenia Grade 2 | 10.48% | 24.15% |
| Neutropenia Grade 3 | 6.03% | 23.67% |
| Thrombocytopenia Grade 2 | 24.76% | 26.09% |
| Thrombocytopenia Grade 3 | 17.62% | 17.39% |

Bone marrow-related adverse events were highly prevalent across all advanced prostate cancer populations. Grade 3 anaemia occurred with high frequency, affecting 48.43% to 52.68% of mHSPC patients and exceeding 60% in the mCRPC cohorts, regardless of treatment modality. Grade 3 neutropenia was prominent in patients undergoing chemotherapy, occurring in 23.22% of the mHSPC cohort and 23.67% of the mCRPC cohort, compared to under 7% in the corresponding NHA arms.

## DISCUSSION

The rising prevalence of advanced prostate cancer in Hong Kong likely reflects population ageing together with increased diagnostic intensity. Consistent with an increasingly chronic disease course, the median duration of systemic treatment in our cohort increased, suggesting a shift toward more sustained multi-line management over time.

Nevertheless, real-world treatment pathways remained largely centered on conventional systemic therapies, namely NHAs and docetaxel, with limited uptake of newer targeted modalities. This finding should be interpreted in the context of potential undercapture of patients receiving care outside local public healthcare systems and the limited availability of certain treatments during earlier observation periods. Consistent with this possibility, when the analysis was restricted to patients diagnosed after 2018, the proportion of patients who received none of the investigated therapies after mHSPC diagnosis decreased from 68% to 47%. Trajectory analyses further demonstrated exceptionally low use of PARP inhibitors, radiopharmaceuticals, and immunotherapies. This pattern contrasts with international guideline recommendations and may reflect local barriers to access, including funding, reimbursement, treatment availability, and institutional capacity.

Across the aggregated cohort, chemotherapy-treated patients had numerically longer median overall survival than those receiving NHAs in both mHSPC (29.2 vs 20.3 months) and mCRPC (14.4 vs 13.3 months). This pattern, however, should be interpreted cautiously. Given the high cost of NHA especially before this option was reimbursed for wider use, systemic chemotherapy may be prioritised for younger, fitter patients with fewer comorbidities and better functional status, whereas oral NHAs are often preferentially used in older or frailer patients who are less suitable for cytotoxic treatment. The apparent survival advantage associated with chemotherapy therefore likely reflects treatment selection and baseline risk differences (“healthy user”/confounding by indication) rather than intrinsic superiority of chemotherapy over NHAs in this observational setting. Patients initially treated with chemotherapy also underwent more subsequent treatment switches. This pattern may reflect their greater fitness to receive additional lines of therapy, as well as clinical treatment sequencing in patients with more advanced or progressive disease.

A major strength of this study is the use of a comprehensive, territory-wide electronic medical record database that captures the majority of care delivered in Hong Kong and enables longitudinal follow-up at the population level. Several limitations merit consideration. First, advanced disease states were ascertained using literature-supported laboratory and diagnostic proxies rather than explicit clinician-recorded staging, which may introduce misclassification. Second, patients managed exclusively in the private sector—particularly those self-funding high-cost novel agents—may be underrepresented in the dataset, which could partly explain the low observed use of targeted therapies.

These findings have important implications for policymakers and health economists by quantifying the contemporary burden of advanced prostate cancer and identifying gaps between recommended and delivered care. Compared with jurisdictions that adopted novel therapies earlier, real-world management in Hong Kong remains constrained by limited later-line options and underuse of innovative treatments, despite their theoretical availability.

Addressing these gaps will require proactive system-level planning as the incidence and survivorship of advanced disease increase. Priorities include sustainable formulary and reimbursement arrangements, molecular testing pathways, and clear referral and treatment algorithms to facilitate the timely and appropriate use of precision therapies. For example, radioligand therapy, particularly ^177^Lu-PSMA-617, is currently positioned after NHAs and taxane chemotherapy or patients are considered appropriate to delay taxane-based chemotherapy in mCRPC. Although its use remains largely limited to later treatment lines, emerging evidence suggests that selected patients may benefit from earlier use in the disease course. Its implementation also requires specialised nuclear medicine facilities, radiopharmaceutical-handling capabilities, and radiopharmaceuticals-enabling policies. Coordinated planning across oncology, nuclear medicine, urology radiology, pharmacy, and hospital administration—supported by transparent health technology assessment and innovative reimbursement mechanisms—could help expand access. Ultimately, aligning patient selection, treatment sequencing, infrastructure readiness, and funding frameworks offers a practical pathway to strengthen precision prostate cancer care in Hong Kong.

## CONCLUSION

This territory-wide study highlights a rapidly escalating advanced prostate cancer burden in Hong Kong. The limited real-world uptake of newer targeted agents reveals critical barriers in precision oncology access. While chemotherapy demonstrated a numerical survival advantage over novel hormonal agents in real-world practice, it incurred a significantly higher burden of severe hematologic toxicities and hospitalisations. These findings deliver crucial population-specific evidence to help local policymakers optimise drug formulary frameworks and improve public healthcare resource allocation for advanced prostate cancer management.

## Data Availability

All code produced in the present study are available upon reasonable request to the authors. The Hong Kong Hospital Authority electronic medical records are confidential, and any request should be approved by Hong Kong Hospital Authority.

